# Match Injury Incidence and Risk Factors in American Youth Soccer: The Elite Clubs National League Injury Data Collection Program

**DOI:** 10.1101/2025.03.14.25323901

**Authors:** Andrew Watson, Jack Behrendt, Ian Staresinic, Jenifer Sanfilippo, Kristin Haraldsdottir

**Affiliations:** From the Departments of Orthopedics and Rehabilitation, University of Wisconsin School of Medicine and Public Health, Madison, WI; The Department of Kinesiology, the University of Wisconsin – Madison; the Division of Intercollegiate Athletics, the University of Wisconsin – Madison

**Author notes:** Address for Correspondence: Andrew Watson, Department of Orthopedics and Rehabilitation, Division of Sports Medicine 1685 Highland Avenue, Madison, WI 53705. **Contributorship Statement:** Drs. Watson and Haraldsdottir conceptualized the study, data collection, data analysis and interpretation, drafted the manuscript, and approved the final manuscript as submitted. Dr. Sanfilippo, Mr. Behrendt and Mr. Staresinic contributed to the data collection, data analysis and interpretation, reviewed the manuscript and approved the final manuscript as submitted.

**Keywords:** adolescent, pediatric, concussion, musculoskeletal

## Abstract

**Objective:** This study aimed to establish a nationwide data collection network in order to identify injury incidence and risk factors during competition in elite youth soccer athletes. We also sought to identify the influence of playing games on consecutive days and the inclusion of a rest day on injury risk in multi-day events.

**Design:** Prospective Cohort Study

**Setting:** Elite Clubs National League (ECNL) events with one game per day over 3 days (regular season, unlimited substitution, no rest days) or 4 days (post-season, limited substitution, rest day between games 1 and 3)

**Participants:** Youth soccer players

**Main Outcome Measures:** Injuries at events were recorded by athletic trainers regarding mechanism, type, body part, return-to-play (yes, no), game number (1-3), and time of year (regular, post-season). Comparisons between groups were made with chi-square tests and the associations of injury incidence with gender, age, game number and time of year were assessed through negative binomial regression models.

**Results:** 806 injuries were reported from 24 events representing 200,047 player-hours. Injury incidence was 4.0 per 1000 player-hours overall and girls had a higher injury incidence than boys (4.4 v 3.5 v per 1,000 player-hours, respectively, incidence rate ratio (IRR)=1.28 [1.0-1.6], p=0.03). Concussion incidence was 0.80 per 1,000 player-hours and did not differ significantly between boys and girls (0.86 v 0.72 per 1,000 player-hours, respectively, IRR=1.28 [0.84-2.0], p=0.26). In the multivariable models, after adjusting for age and gender, injury incidence was not associated with game number (game 2 IRR=0.87 [0.73-1.0], p=0.12; game 3 IRR=0.87 [0.73-1.0], p=0.13). No significant interactions between game number and time of year (rest day or not) were identified for all injuries (Game [3] * Time (regular season) IRR=1.16 [0.65-2.1], p=0.61) or concussions only (1.25 [0.3-5.2], p=0.76).

**Conclusions:** Among this group of elite youth soccer players, girls had a higher overall injury incidence, but concussion incidence was similar. No increased injury incidence was identified during the later games of multi-day events when games were played on consecutive days with unlimited substitution, and injury incidence was not influenced by the inclusion of rest days. The establishment of this novel data collection framework embedded within a nationwide elite youth soccer organization will continue to inform strategies to promote safe and healthy participation in youth soccer events.

**Key Points:**

- The incorporation of athletic trainers at youth soccer events represents an opportunity to develop a novel, nationwide data collection network to evaluate match injury characteristics, incidence and risk factors.
- Female youth soccer athletes had a higher overall injury incidence, although no difference was identified in injury mechanism, severity, or concussion incidence.
- With unlimited substitution, playing 3 full-length games on 3 consecutive days was not associated with an increased injury or concussion incidence in later games, and the incorporation of rest days in multi-day events did not appear to influence injury or concussion risk.

## INTRODUCTION

Although youth sports have been shown to have significant positive physical and mental health impacts for children, sport participation carries an inherent risk of injury. Injuries among youth soccer athletes occur primarily through contact, predominantly involving the lower extremities, and may be increasing over the last several decades.^1–3^ Importantly, prior research has consistently found that injuries are far more likely in matches than in training.^1,4,5^ Therefore, efforts to identify risk factors for match injuries represent an important opportunity to guide broad interventions within youth soccer to reduce injury risk.

Although various risk factors have been identified for youth soccer injuries, prior work has primarily focused on individual-level risk factors and specific types of injuries such as anterior cruciate ligament (ACL) injuries and concussions.^5–8^ For example, research has found that individuals with greater knee valgus are at an increased risk of ACL injury, leading to the development of neuromuscular training programs that have been found to reduce the risk of ACL and lower extremity injuries.^9–12^ Less prior work, however, has focused on how policies or operational-level decisions can impact injury risk. For example, fatigue has been identified as a risk factor for injury, and it has been suggested that accumulated fatigue over multiple days of competition or multiple games in a single day could lead to an increased risk of injury.^13^ We are aware of no prior research that has attempted to quantify this proposed increased incidence and whether this association is influenced by characteristics like age or gender.

Within collegiate athletes, the establishment of the NCAA Injury Surveillance Program has been used to quantify the incidence of injuries and identify sport and sex-specific risk factors to guide interventions.^14^ Similar data collection frameworks in high school sports have been used to evaluate the epidemiology of certain injuries, associated risk factors, and interventions within certain populations.^15–18^ American youth club sports represent a different context than high school or collegiate sports, however, and we are aware of no prior efforts to develop an injury data collection framework embedded within a nationwide youth sports organization. The establishment of a longitudinal injury monitoring program in youth sports can help identify generalizable, modifiable risk factors for injury as well as inform and evaluate operational-level interventions to reduce risk.

The Elite Clubs National League (ECNL) is a nationwide youth soccer competition and coach development platform. In addition to hosting regional conference-based competitions, the ECNL hosts national events throughout the country at various times of the year, as well as post-season competitions. National events consist of around 100-200 teams playing one game per day in a single location over 3 days. Post-season competition durations differ based on the number of teams but initially consist of 3 games played over 4 days, with a rest day incorporated either between games 1 and 2 or games 2 and 3. The purpose of this study was to establish an injury data collection framework through the embedded athletic trainers at ECNL events in order to identify the incidence of injuries requiring medical attention during youth soccer matches, as well as the associations with various risk factors and their interactions. We hypothesized that 1) female and older athletes would have a higher rate of injuries overall, as well as concussions, 2) injury incidence would increase on the third game of events, and 3) the inclusion of a rest day would be associated with a reduced increase in injury incidence from the first to the third game.

## METHODS

### Study Design

All procedures performed in this study were approved by the Institutional Review Board of the University of Wisconsin-Madison. At ECNL events from October 2023 through June 2024, licensed athletic trainers (LATs) evaluated and documented all injuries presenting to medical tents distributed throughout the competition facility. For each participant interaction, LATs recorded the date, event location, participant age group (U13, U14, U15, U16, U17 or U18) and gender, as well as the injury date (if different than the current date), mechanism (contact, non-contact), body part involved, type of injury (sprain, strain, concussion, contusion, fracture, laceration, dislocation, other), the treatment provided and whether the athlete was restricted from play (yes, no). The de-identified injury reports were provided to the study team after the conclusion of the event. The game number (1, 2, or 3) was determined based on the date of injury and age group as all teams from the same age group participate on the same day.

Regular season events consisted of 3 full-length games with unlimited substitution played on 3 consecutive days. Post-season events consisted of 3 full-length games with no re-entry during the same half played over 4 days, such that a rest day was included between either games 1 and 2 or games 3 and 4.

### Statistical Analysis

Data were initially evaluated using descriptive statistics, including estimates of central tendency (mean, median) and variability (standard deviation, interquartile range, range) for continuous variables, and counts and percentages for categorical variables. Injury characteristics (body part involved, injury type, mechanism, and return to play) were compared between genders and age groups with chi-square tests. Injury incidence was expressed as the number of total reported injuries per 1,000 player-hours overall and separately for suspected concussions per 1,000 player-hours for boys and girls. Player-hours for each day of each event were calculated separately for each age group and gender based on the number of games played, game duration (minutes) and number of players on the field. For example, U18 boys’ games are 90 minutes long with 22 players on the field at once. If fifty U18 boys’ games were played on a specific day, this would result in 1,650 player-hours (50 games x 90 minutes/game x 22 players ÷ 60 minutes/hour). Confidence intervals for injury incidence were calculated using the exact method.^19^

Injury incidence was compared between genders and age groups using incidence rate ratios (IRR) from negative binomial regressions to predict injury with the log of player-days as an offset for 1) all injuries and 2) concussions. Similar negative binomial regression models were developed to compare IRRs between regular and post-season, adjusted for age group and gender. To evaluate the influence of game number on injury incidence, similar multivariable negative binomial regression models were developed to predict the number of injuries, with age group, gender, game number, and the log of player-days as an offset for 1) all injuries and 2) concussions. To evaluate the potential influence of rest days, separate negative binomial regression models were developed to predict the number of injuries, including the interaction of game number (1, 2, or 3) and time of year (regular season, post-season) and the log of player-hours as an offset. Coefficients from negative binomial models were exponentiated to represent IRRs for binary variables and Wald confidence intervals were generated. The significance level was determined *a priori* at the 0.05 level and all tests were 2-tailed. All statistical analyses were performed in R.

## RESULTS

Data was collected from 24 events, representing over 200,000 player-hours and including 802 injuries, of which 160 were concussions. Head (28%), knee (19%), and ankle (18%) were the most commonly injured body parts, and sprains (32%), concussions (20%), and contusions (18%) were the most common injury types (See Table 1). The majority of injuries were due to a contact mechanism with no difference between genders (see Table 2). Similarly, the majority of injuries resulted in restriction from play with no difference between genders (see Table 3).

**Table 1.** The distribution of body parts and injury types for match injuries in youth soccer athletes.

|  | All Athletes | Boys | Girls | p<sup>a</sup> |
| --- | --- | --- | --- | --- |
| Body Part |  |  |  | 0.032 |
| Head | 221 (28%) | 90 (29%) | 131 (27%) |  |
| Knee | 153 (19%) | 36 (12%) | 117 (24%) |  |
| Ankle | 145 (18%) | 60 (20%) | 85 (17%) |  |
| Arm/Hand | 54 (6.7%) | 25 (8.1%) | 29 (5.9%) |  |
| Face | 53 (6.6%) | 19 (6.2%) | 34 (6.9%) |  |
| Shoulder | 39 (4.9%) | 17 (5.5%) | 22 (4.5%) |  |
| Hip | 32 (4%) | 15 (4.9%) | 17 (3.4%) |  |
| Upper Leg | 32 (4%) | 17 (5.5%) | 15 (3%) |  |
| Trunk | 26 (3.2%) | 10 (3.3%) | 16 (3.2%) |  |
| Lower Leg | 23 (2.9%) | 8 (2.6%) | 15 (3%) |  |
| Foot | 20 (2.5%) | 9 (2.9%) | 11 (2.2%) |  |
| Other | 2 (0.25%) | 1 (0.33%) | 1 (0.2%) |  |
| Injury Type |  |  |  | 0.012 |
| Sprain | 255 (32%) | 89 (29%) | 166 (34%) |  |
| Concussion | 160 (20%) | 63 (21%) | 97 (20%) |  |
| Contusion | 147 (18%) | 46 (15%) | 101 (20%) |  |
| Fracture | 100 (12%) | 46 (15%) | 54 (11%) |  |
| Strain | 73 (9.1%) | 38 (12%) | 35 (7.1%) |  |
| Laceration | 30 (3.7%) | 15 (4.9%) | 15 (3%) |  |
| Dislocation | 19 (2.4%) | 6 (2%) | 13 (2.6%) |  |
| Other | 17 (2.1%) | 3 (0.98%) | 14 (2.8%) |  |

Data presented as N (%). <sup>a</sup>Comparison between genders made with chi-square tests.

**Table 2.** Injury mechanism and severity by gender and age group in youth soccer players.

|  | <u>Mechanism</u> |  | <u>Severity</u> |  |
| --- | --- | --- | --- | --- |
|  | Contact | p | Restriction from play | p |
| All athletes | 600 (74.8%) |  | 606 (75.6%) |  |
| Gender |  | 0.76 |  | 0.80 |
| Girls | 368 (74.3%) |  | 372 (75.2%) |  |
| Boys | 232 (75.6%) |  | 234 (76.2%) |  |
| Age group |  | 0.46 |  | 0.005 |
| U13 | 44 (81.5%) |  | 37 (68.5%) |  |
| U14 | 60 (73.2%) |  | 66 (80.5%) |  |
| U15 | 90 (68.7%) |  | 84 (64.1%) |  |
| U16 | 165 (77.1%) |  | 175 (81.8%) |  |
| U17 | 144 (75.4%) |  | 148 (77.5%) |  |
| U18 | 97 (74.6%) |  | 96 (73.8%) |  |

Data presented as N (%). <sup>a</sup>Comparisons made between genders and age groups with chi-square tests.

**Table 3.** Total match injuries, player-hours and injury incidence among youth soccer athletes by gender, age group and time of year.

| Group | Total injuries | Concussions | Player-hours | Injury incidence<sup>a</sup> | Concussion incidence<sup>a</sup> |
| --- | --- | --- | --- | --- | --- |
| Gender |  |  |  |  |  |
| Boys | 307 | 63 | 87,092 | 3.5 | 0.72 |
| Girls | 495 | 97 | 112,955 | 4.4<sup>b</sup> | 0.86 |
| Age group |  |  |  |  |  |
| U13 | 54 | 9 | 17,158 | 3.2 | 0.53 |
| U14 | 82 | 17 | 18,454 | 4.4 | 0.92 |
| U15 | 131 | 34 | 27,368 | 4.8<sup>c</sup> | 1.2<sup>c</sup> |
| U16 | 214 | 45 | 42,357 | 5.1<sup>c</sup> | 1.1 |
| U17 | 191 | 30 | 54,434 | 3.5 | 0.55 |
| U18 | 130 | 25 | 40,276 | 3.2 | 0.62 |
| Time of year |  |  |  |  |  |
| Regular Season | 588 | 131 | 143,434 | 4.1 | 0.91 |
| Post-season | 214 | 29 | 56,613 | 3.8 | 0.51<sup>d</sup> |
| Total | 802 | 160 | 200,047 | 4.0 | 0.80 |

<sup>a</sup>Incidence expressed as 1,000 player-hours; <sup>b</sup>p<0.05 for incidence rate ratio v boys; <sup>c</sup>p<0.05 for
incidence rate ratio v U13 (reference); <sup>c</sup>p<0.05 for incidence rate ratio v regular season.

The number of injuries, player-hours, and injury incidence for all injuries and concussions by gender, age group and time of year are shown in Table 3. Overall injury incidence was higher for girls (IRR = 1.28 [95% CI = 1.0-1.6], p=0.03) but the difference in concussion incidence was not statistically significant (1.28 [0.84-2.0], p=0.26). Compared to U13 as the reference age group, U15 and U16 players had a higher overall injury incidence (U15 IRR = 1.51 [1.0-2.2], p=0.038; U16 IRR=1.55 [1.1-2.3], p=0.021) and U15 players had a higher concussion incidence (IRR = 2.34 [1.1-5.6], p=0.039). After adjusting for age group and gender, injury incidence in the regular season was not significantly different than the post-season (1.08 [0.93-1.27], p=0.33), although concussion incidence was higher in the regular season (1.78 [1.19-2.77], p=0.004).

The unadjusted injury incidence by game number is shown in Figure 1. For all injuries, after adjusting for age group and gender, no significant independent associations were identified between injury incidence and game number (game 2=0.89 [0.66-1.2], p=0.38; game 3=0.86 [0.65-1.1], p=0.29). Similarly, after adjusting for age group and gender, no significant independent associations were identified between concussion incidence and game number (game 2=0.97 [0.57-1.6], p=0.91; game 3=1.07 [0.64-1.08], p=0.78). The adjusted IRRs by game number for all injuries and concussions are shown in Figure 2. The injury incidence by game number and time of year is shown in Figure 3. No significant interactions between game number and time of year (incorporated rest day in post-season only) were identified for all injuries (Game [3] * Time (regular season) IRR=1.16 [0.65-2.1], p=0.61) or concussions (1.25 [0.3-5.2], p=0.76).

**Figure 1.**
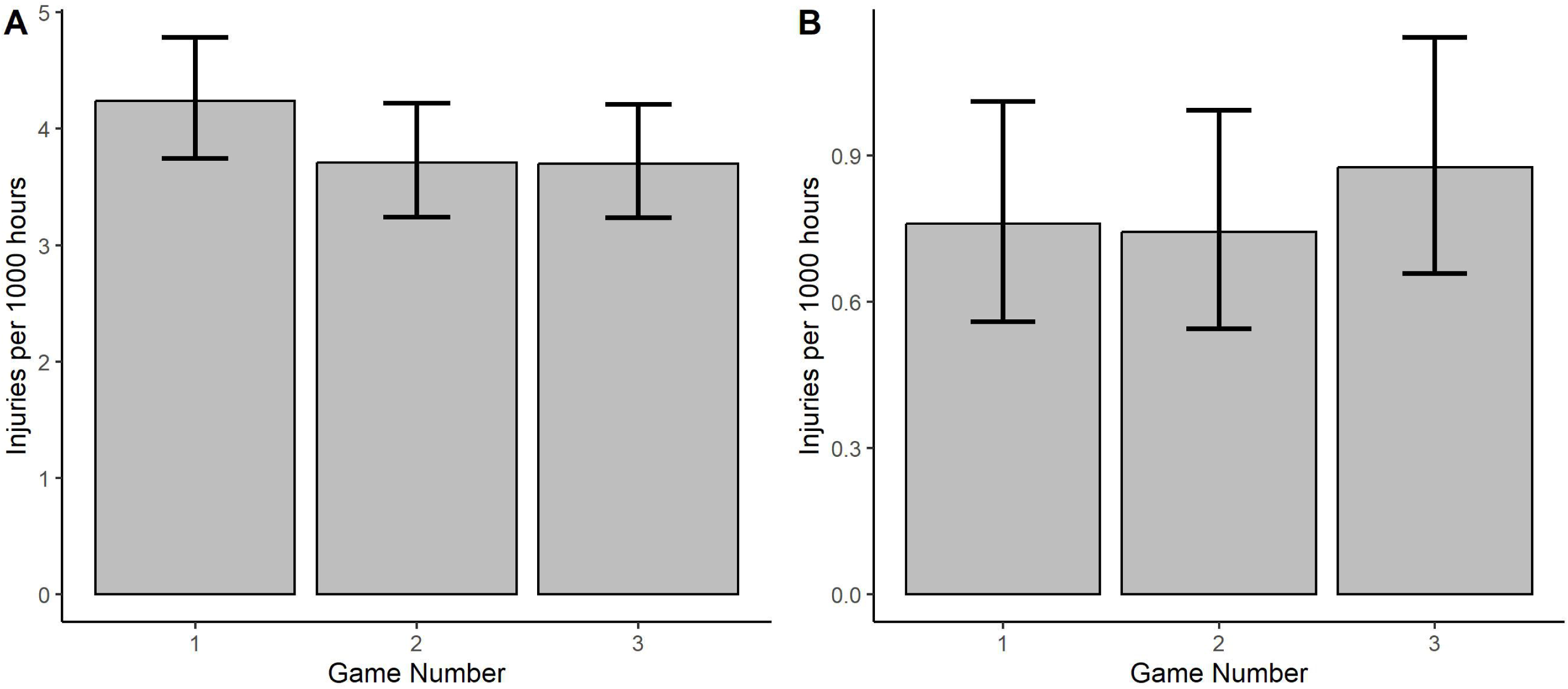
Match injury incidence by game of event among youth soccer athletes for (A) all injuries and (B) concussions.

**Figure 2.**
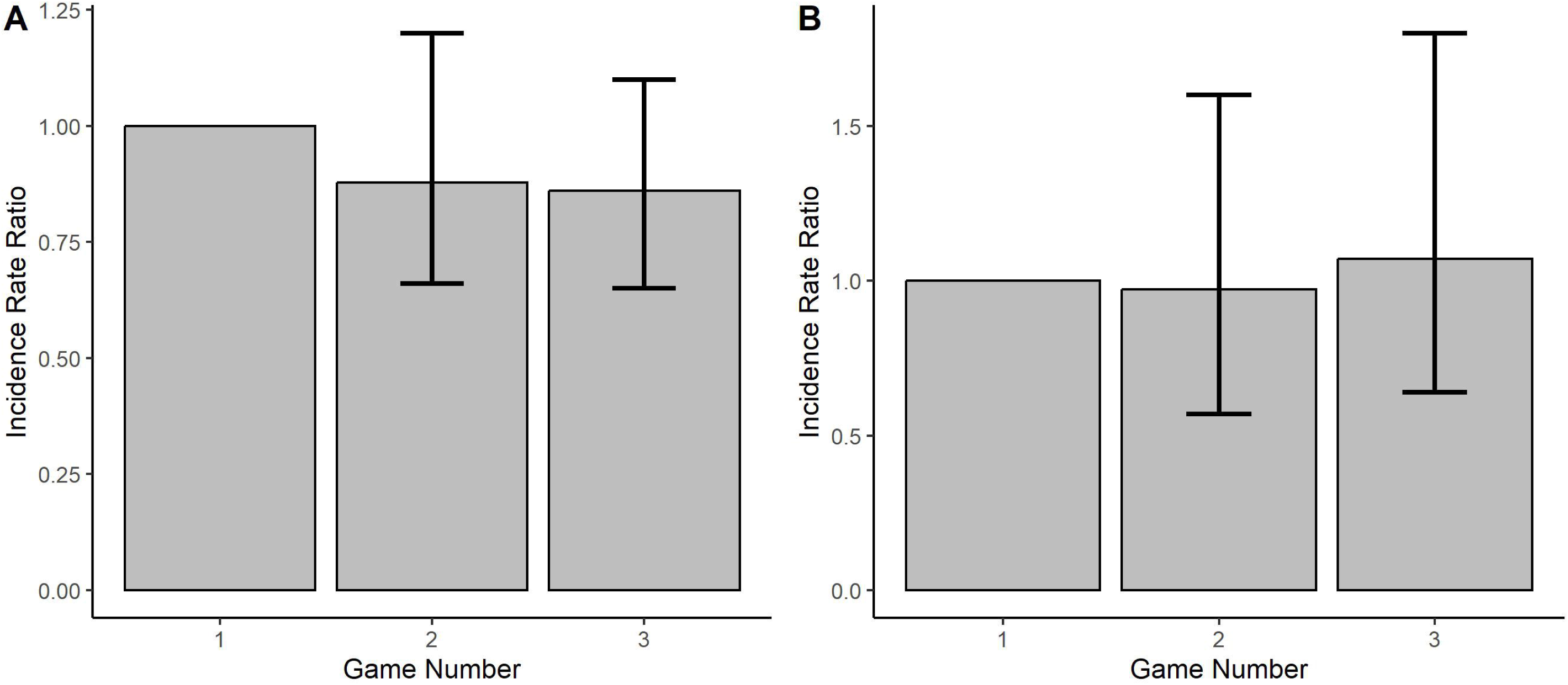
Match injury rate ratios by game of event in youth soccer athletes (with game 1 as reference), adjusted for age and gender for (A) all injuries and (B) concussions.

**Figure 3.**
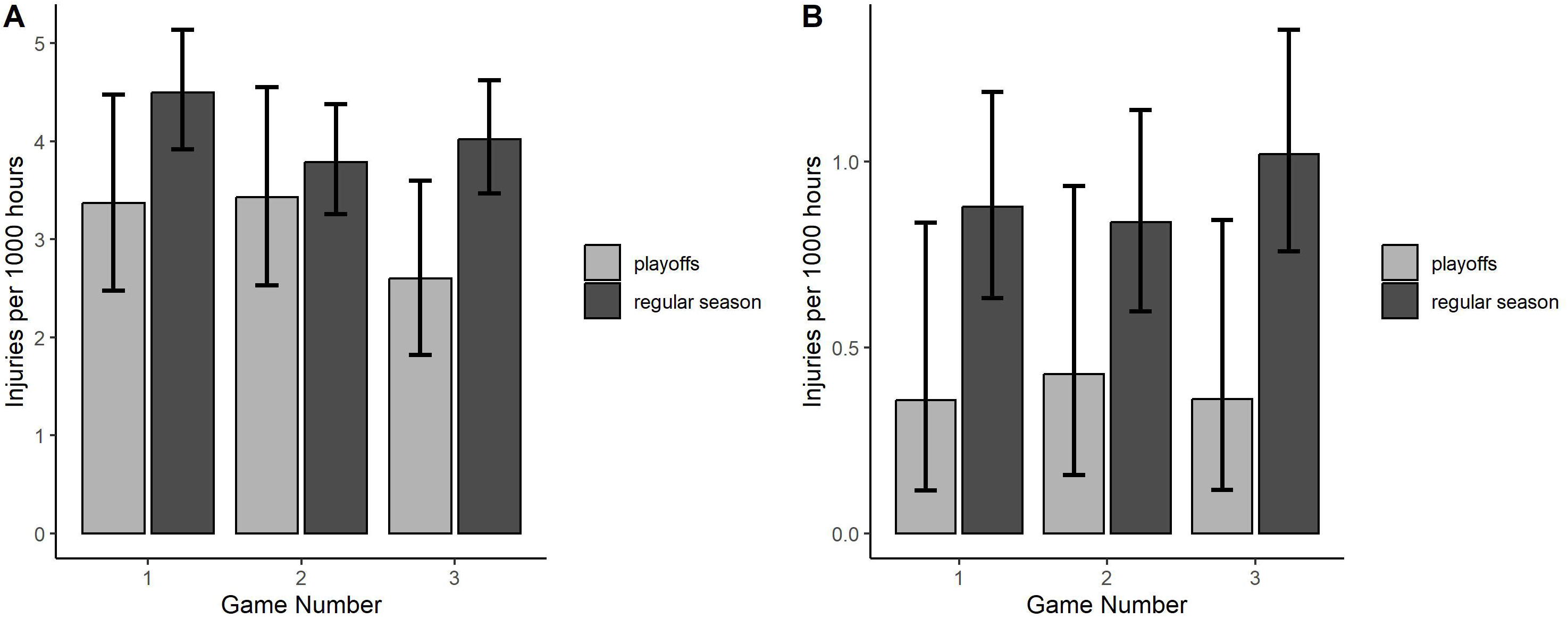
Injury incidence by game of event and time of year in youth soccer athletes for (A) all injuries and (B) concussions.

## DISCUSSION

This study represents the initial results from a novel youth soccer injury data collection framework utilizing licensed athletic trainers operating within multi-day events throughout the United States. This unique program can help to better define injury mechanisms, incidence and risk factors within youth soccer to inform policies and evaluate their impact on injuries during competition. Although we did identify a higher overall injury incidence among female athletes consistent with prior research,^3,4^ the higher incidence of concussions in female athletes was not statistically significant. We also found that U15 and U16 age groups had a higher overall injury incidence and U15 athletes had a higher concussion incidence than other age groups. The overall injury incidence identified here was lower than that reported in prior research of youth soccer match injuries,^4,20–22^ although this may be partly attributable to differences in data collection methods and the fact that in the present study, minor injuries were less likely to be evaluated by the athletic trainers at ECNL events than may have been captured under other data collection procedures. Nonetheless, the incidence of more serious injuries such as concussions, which would be expected to be evaluated at these events, was still slightly lower than previously published data from youth soccer matches.^23,24^

We found that although overall injuries did not differ significantly by time of year, concussion incidence was lower in the post-season than the regular season. The underlying reason for this is not immediately clear and we are not familiar with prior work that has evaluated this relationship. Higher physical fitness has been found to be associated with a reduced risk of injury in both youth and collegiate soccer players,^25,26^ and higher levels of fitness at the end of the season may offer a protective effect. Fair play, the expectation that athletes abide by the laws of the game, has been suggested as a protective factor for injury,^27,28^ and players in the post-season may be more incentivized to avoid reckless or illegal play given the increased stakes at that time of year. This is speculative, however, and the underlying mechanisms for this relationship remain an important area of future research.

Injury incidence decreased slightly from game 1 to game 3 of events in both the regular season and post-season, suggesting that any accumulated fatigue from this competition format does not lead to an increased risk of injury in later games. It has been suggested that participating in multiple games in the same day can increase the risk of injury, but no prior research has evaluated the change in injury incidence in youth soccer when single, full-length games are played on consecutive days. In this study, we found that playing three games on consecutive days with unlimited substitution was not associated with an increased injury risk during games two or three. Similarly, we did not find any evidence that intervening rest days in the post-season influence the injury risk in the later games of events. Although prior research has suggested that fatigue may play a role in injury risk,^29^ particularly during periods of match congestion,^30^ these findings suggest that there is no appreciable increase in risk throughout 3-4 day events when participation is limited to no more than one game per day. It also suggests that there is no significant influence of the incorporation of a rest day within the context of a 3-game event with respect to injury risk. It is possible that the increased rest may influence the quality of the games and other individual-level outcomes (soreness, fatigue, e.g.), but these are not evaluated here and are beyond the scope of the current study.

Importantly, this research represents the first available evidence from the establishment of a novel data collection program embedded within the operations of a nationwide youth soccer competition platform. Longitudinal data collection within this framework can continue to evaluate additional risk factors (weather, playing surface, e.g.) and how these interact with individual characteristics to influence injury on a scale that is generalizable to a broad population of youth athletes. Similar to prior efforts within collegiate sports,^14^ it also serves as an innovative opportunity to facilitate data-driven operational decisions and policies regarding player safety, and evaluate the impacts of these changes on subsequent injury risk. We are aware of no prior longitudinal injury surveillance programs on this scale within youth sports in the United States.

This study has several limitations. The information collected by the athletic trainers is relatively limited due to the time constraints of patient interactions and the efforts to maintain confidentiality for those athletes involved. Importantly, any injured athletes who did not seek care at a medical tent at the event would not be captured and this may explain differences in injury incidence in this study when compared to injuries collected through different mechanisms in prior work. Nonetheless, in order for this to undermine the associations identified between injury incidence and risk factors within this sample, any potential confounding factor would have to be related to both the likelihood to seek care and the factor being evaluated. We have no reason to suspect that the likelihood to seek care would be related to gender, age, or time of year or but we cannot account for this possibility. It is also possible that risk factors not accounted for in this analysis could be related to both injury incidence and the risk factors analyzed, potentially confounding the results. Finally, while this data represents information regarding a large number of male and female soccer athletes from a nationwide sample, it is unknown whether this sample is sufficiently heterogeneous with respect to characteristics such as geography or socioeconomic status to be generalizable to all youth soccer athletes. In addition, it may not be generalizable to older athlete populations or other sports.

In conclusion, this study represents the initial analysis of injury incidence rates and risk factors for youth soccer match injuries from an injury surveillance program embedded within a nationwide competitive platform. Overall injury incidence was higher in girls, but concussion incidence was similar between genders. We found that injury incidence did not increase in the later games of events consisting of 3 games with unlimited substitution on 3 consecutive days. While we did identify lower concussion incidence in the post-season compared to the regular season, the inclusion of rest days between games 1 and 3 in the post-season did not appear to influence overall injury or concussion incidence in the later stages of the events. The establishment of this novel, longitudinal data collection framework can help guide future research to identify at-risk groups for specific types of injuries and modifiable risk factors that can inform and subsequently evaluate operational procedures and policies to improve player safety.

## Data Availability

All data produced in the present study are available upon reasonable request to the authors

## ACKNOWLEDGMENTS

There are no funding sources to report for this study. We are extremely grateful to the staff, coaches, players and families within the ECNL that made this research possible.

